# Gait-to-Contact (G2C) - A novel deep learning framework to predict total knee replacement wear from gait patterns

**DOI:** 10.1101/2024.09.27.24314383

**Authors:** Mattia Perrone, Scott Simmons, Philip Malloy, Catherine Yuh, John Martin, Steven P. Mell

## Abstract

**Background:** Total knee replacement (TKR) is the most common inpatient surgery in the US. Studies leveraging finite element analysis (FEA) models have shown that variability of gait patterns can lead to significant variability of wear rates in TKR settings. However, FEA models can be resource-intensive and time-consuming to execute, hindering further research in this area. This study introduces a novel deep learning-based surrogate modeling approach aimed at significantly reducing computational costs and processing time compared to traditional FEA models.

**Methods:** A published method was used to generate 314 variations of ISO14243-3(2014) anterior/posterior translation, internal/external rotation, flexion/extension, and axial loading time series, and a validated FEA model was used to calculate linear wear distribution on the polyethylene liner. A deep learning model featuring a transformer-CNN based encoder-decoder architecture was trained to predict linear wear distribution using gait pattern time series as input. Model performance was evaluated by comparing the deep learning and FEA model predictions using metrics such as mean absolute percentage error (MAPE) for relevant geometric features of the wear scar, structural similarity index measure (SSIM) and normalized mutual information (NMI).

**Results:** The deep learning model significantly reduced the computational time for generating wear predictions compared to FEA, with the former training and inferring in minutes, and the latter requiring days. Comparisons of deep learning model wear map predictions to FEA results yielded MAPE values below 6% for most of the variables and SSIM and NMI values above 0.88, indicating a high level of agreement.

**Conclusion:** The deep learning approach provides a promising alternative to FEA for predicting wear in TKR, with substantial reductions in computational time and comparable accuracy. Future research will aim to apply this methodology to clinical patient data, which could lead to more personalized and timely interventions in TKR settings.

## Introduction

Total knee replacement (TKR) is the most common inpatient surgery in the US^1^, with annual procedures anticipated to increase to 3.48 million by 2030^2^. It is estimated that over 4.2% of adults are living with a TKR in the US alone^3,4^. Additionally, there is a demographic trend towards younger and more active patients undergoing TKR surgery, raising the concern of component wear^5,6^. The growing number of individuals undergoing TKR surgeries magnifies the economic and societal burden of even a small percentage of failures, and underscores the importance of comprehending the causes behind the failure of TKR. Wear of ultra-high molecular weight polyethylene and the body’s reaction to the resulting debris are among the most significant factors for TKR revision^7,8,9^, especially in cases of late failure. Although advancements in materials have significantly lowered the rates of complications due to wear, an enhanced understanding of the patient-specific factors that contribute to wear could further decrease these rates^10^. One of the most critical patient-specific factors known to contribute to excess wear is the patient’s movement patterns during walking, in particular knee kinematics and kinetics^11,12^. Identifying how aberrant knee kinematics and kinetics contribute to wear may allow for early intervention in high-risk patients.

Finite element analysis (FEA) is a computational tool extensively used to investigate wear profiles in TKR^12,13^, and has demonstrated that variability in gait can lead to variability in implant wear^10^. However, FEA can be resource-intensive and time-consuming to execute, with many studies reporting a computational time of hours or even days^14^. This may hinder further large-scale parametric investigations, prompting the exploration of alternative methodologies to reduce processing time. Surrogate modeling is an approach where a computationally efficient mathematical approximation of a system is built using data selectively generated from a more complex and high-fidelity simulation such as FEA^15^. We have previously used this approach to create surrogate linear regression models to predict wear rates in TKRs under a variety of conditions^13,16,10^. However, these models only predict volumetric wear rates, which neglect the spatial distribution of wear provided by FEA models. This spatial information may be important for understanding failure. For example, a small but deep wear scar may be more detrimental than a more diffuse wear scar, despite both having the same overall wear volume. Likewise, a wear scar situated posteriorly, or with a greater angle between the medial and lateral compartments, may demonstrate to clinicians that implant malalignment is the source of wear debris and a subsequent foreign body reaction.

Recently, growing interest in deep learning has increased its application within the computational biomechanics domain. Several studies have used deep learning models as surrogate models, presenting a promising alternative to conventional FEA approaches, with a notable advantage in terms of processing time^14^. Such surrogate models have found applications in various areas, including studies of the liver^17^, breast^18^, and thoracic aorta^19^. Most of these studies concentrate on assessing deformations in anatomical regions, which is a common output of FEA. However, there is a limited number of investigations employing this methodology in analysis of total joint replacements, with none aimed at predicting mechanical wear in total knee implants. While some studies have attempted to use surrogate machine learning models to predict strains^20^ and stresses^21^ at the joint interface, they have not employed deep learning architectures to predict spatial relationships when extracting features, such as convolutional or graph neural networks. Furthermore, no studies have utilized time series data such as gait kinematics and kinetics to predict surface wear patterns.

In this study, we introduce a deep learning framework capable of predicting tibial polyethylene wear in a TKR starting from kinematic and kinetic gait patterns, lowering computational costs and processing time compared to FEA. We utilized an existing dataset of 314 FEA simulations where kinematics and kinetics were modified to predict polyethylene wear in TKRs. Unlike previous studies that utilize fully-connected neural networks or traditional machine learning algorithms, our approach introduces a novel transformer encoder paired to a CNN decoder architecture, and predict spatial polyethylene wear from gait. This architecture enables the generation of polyethylene wear maps as output data to visually assess implant performance in a clinical setting. To our knowledge, this is the first surrogate deep learning model for FEA in TKR settings.

## Methods

### Training dataset

In a previous study we generated 314 unique variations of gait standard ISO14243-3(2014), including anterior/posterior translation, internal/external rotation, flexion/extension, and axial loading^10^, using Latin Hypercube Sampling^22^. These unique waveforms were used as inputs to a previously published and validated TKR model^10,13,16,23^, which was used to calculate linear wear distribution on the polyethylene liner. All time series generated by the previous study were normalized to 100 time steps, resulting in a total shape of 314×100×4 for our model input data. For our ground truth model output, the mesh of the polyethylene tibial insert was exported from Abaqus v2017/Standard (Dassault Systèmes, Waltham, MA) and interpolated to a 100 × 100 pixel grid using Python (version 3.12.1), so that each pixel represents a specific value of linear wear.

### Model architecture

A deep learning model was developed to predict polyethylene wear on the interpolated image starting from the gait pattern time series (**Figure 1**). We implemented an encoder-decoder architecture incorporating a dual-head transformer encoder and transposed convolutional layers in the decoder. Our model uses the transformer encoder to condense gait time-series into a compressed but feature-rich latent space, while the decoder employs transposed convolutional layers to reconstruct pixel-wise spatial structure from latent feature representations. We specifically chose to use transformers, as they are often better able to manage long-range dependencies within the time series data compared to other commonly used models such as long short-term memory models and recurrent neural networks^24^. In designing the model architecture, we thoroughly tuned model hyperparameters on the transformer encoder to enhance feature learning^25^ and experimented with the depths and widths of decoder layers.

**Figure 1:**
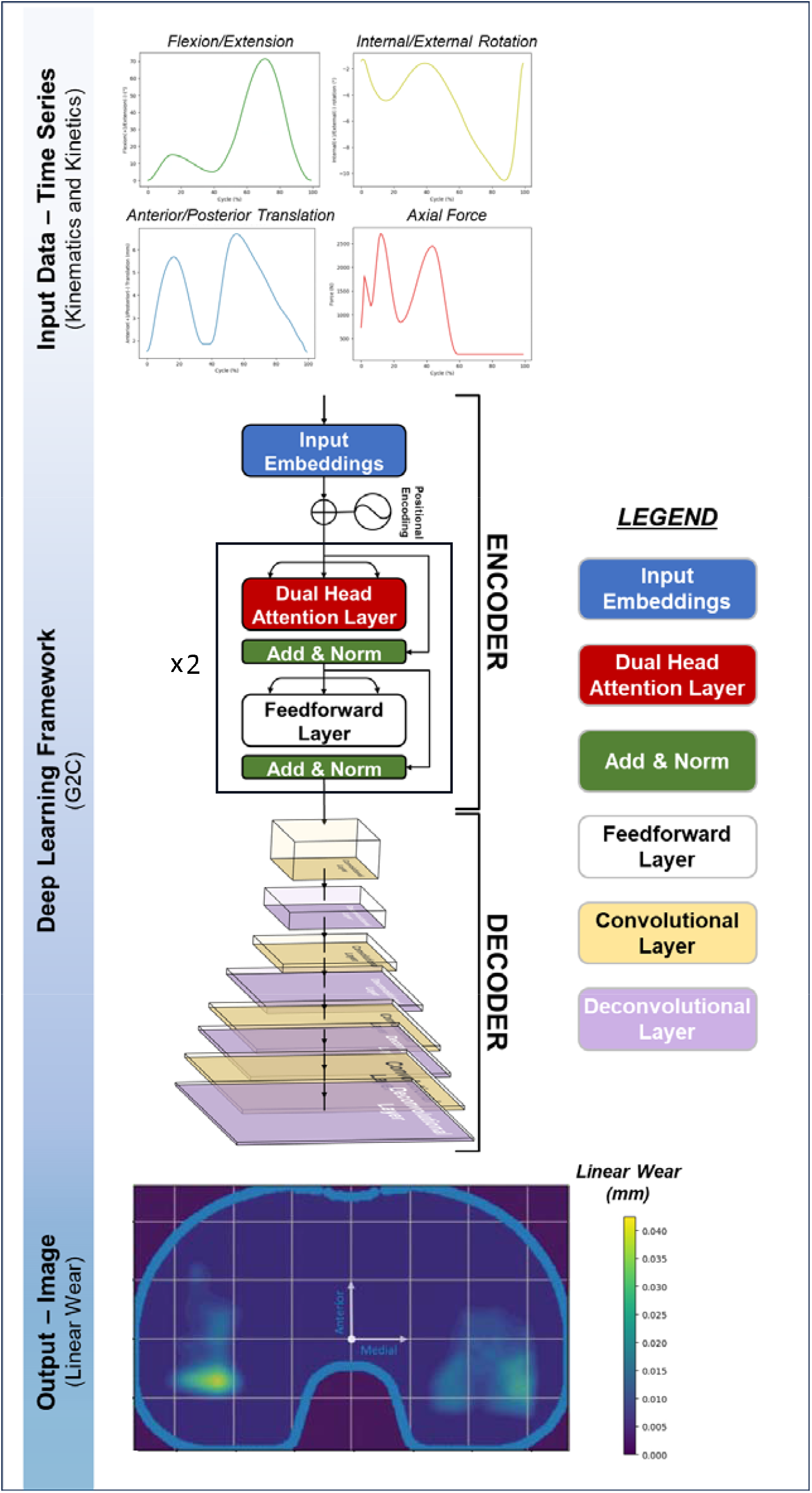
The proposed model consists of a U-net architecture incorporating a dual-head transformer encoder to discern temporal dependencies within the multivariate time series and deconvolutional layers in the decoder to ensure the reconstruction of image data. 314 input timeseries were normalized to 100 points, resulting in a total shape of 314×100×4. The model outputs a 100×100 pixel linear wear map prediction.

For the decoder, we employed transposed convolutional layers to reconstruct the spatial dimensions from the compressed representations and convolution layers to enhance feature representations by extracting spatial details. Specifically, we leveraged a strategy of progressively halving the channel dimensions for each transposed convolutional layer. This setup of combining convolutional and transposed convolutional layers is particularly beneficial in tasks requiring generation of high-resolution outputs from lower-resolution inputs^26^. This approach, combined with the use of smaller (3) and larger (6) kernel sizes for transposed convolutional and convolutional layers, respectively, allows for an optimal trade-off between capturing local details and aggregating broader contextual information. Strides of 2 across all deconvolutional layers ensure effective dimensionality reduction and expansion without excessive computational costs. The final layer of the decoder uses a 1×1 convolution to transform the multi-channel feature maps into the desired single-channel output consisting of the wear value for each specific pixel.

### Model training and evaluation metrics

Hyperparameters tested in the encoder include the input dimension, the number of transformer layers and transformer attention heads, the number of feedforward layers within each transformer layer, as well as training parameters (e.g., learning rate) (**Table 1**). The optimal configuration identified for these hyperparameters were an input dimension of 128, 2 transformer heads, 2 transformer layers each with a feedforward layer of 256, and a learning rate of 9.5 × 10^-5^. To identify the optimal configuration across a broad range of hyperparameters, we employed Optuna^27^, an optimization framework designed to automate hyperparameter tuning (version 3.6.1). Specifically, tree-structured Parzen estimators^28^ (TPE) were used to model the probability distributions of hyperparameters and guide the search towards the most effective configurations.

**Table 1:**
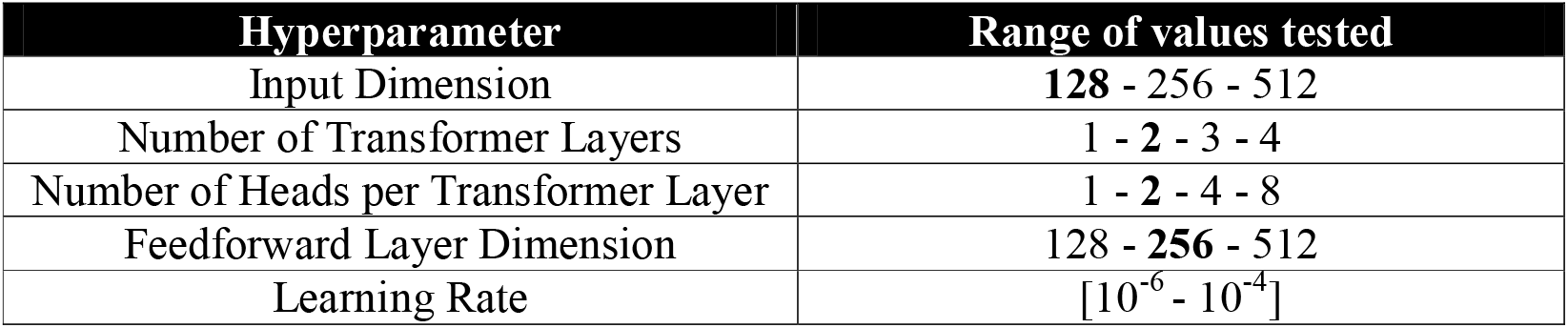
Overview of the range of values tested for each hyperparameter in the study. The bolded values indicate the hyperparameters that were selected for the final model configuration. Square brackets denote that any value within the specified extremes was eligible for testing.

| Hyperparameter | Range of values tested |
| --- | --- |
| Input Dimension | **128** - 256 - 512 |
| Number of Transformer Layers | 1 - **2** - 3 - 4 |
| Number of Heads per Transformer Layer | 1 - **2** - 4 - 8 |
| Feedforward Layer Dimension | 128 - **256** - 512 |
| Learning Rate | $[10^{-6} - 10^{-4}]$ |

Before training the model, each input feature was individually normalized to a range of 0 to 1 to improve model predictions. After performing a random 60/20/20 training/validation/test split, we trained the model using mean squared error as the loss function, conducted hyperparameter tuning on the validation set and assessed the performance of the best model on the test set. Comparisons were made between predictions from the deep learning model and the gold standard predictions from the FEA model. These were assessed in both the medial and lateral compartments, using the mean absolute percentage error (MAPE) as error metric. MAPE is defined as follows:

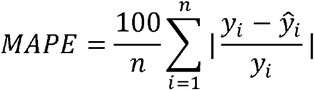

where *ŷ* are the predictions from the deep learning model, *y* are the ground truth from the FEA model and n is the total number of samples in the dataset. To provide a comprehensive assessment of the predicted wear scar heatmaps, we examined metrics that are particularly relevant to the evaluation of wear scars in TKR settings, along with those commonly used in general heatmap analysis. Wear scar area (weighted and non-weighted) and angle between centroids were evaluated for their specific relevance to TKR wear patterns^16,29,30,31^ (**Figure 2**), where weighted wear scar area was determined by summing the values of linear wear across all pixels, while non weighted wear scar area was calculated by identifying pixels that fall within the wear scar boundary and simply summing their count. NMI (Normalized Mutual Information) and SSIM (Structural Similarity Index Measure) were utilized for broader pattern identification across heatmaps^32,33^. Specifically, SSIM assessed the spatial fidelity between FEA and deep learning heatmaps, while NMI measured the shared information between the heatmaps predicted by these two methodologies. We also investigated other metrics, such as width and length of the wear scar (**Figure 2**). All these parameters were evaluated both for the medial and lateral compartments of the implant. To assess whether the grid size used for interpolation of the wear scar influenced the results when exporting the mesh from Abaqus, we analyzed different configurations with grid sizes of 100×100, 150×150, and 200×200 pixels. Python libraries used for data preprocessing, model implementation and hyperparameters tuning include Pandas, Numpy, Scipy, Scikit-learn, PyTorch, PyTorch Lightning, and Optuna. The model was trained on an NVIDIA A5000 GPU with 24 GB of VRAM.

**Figure 2:**
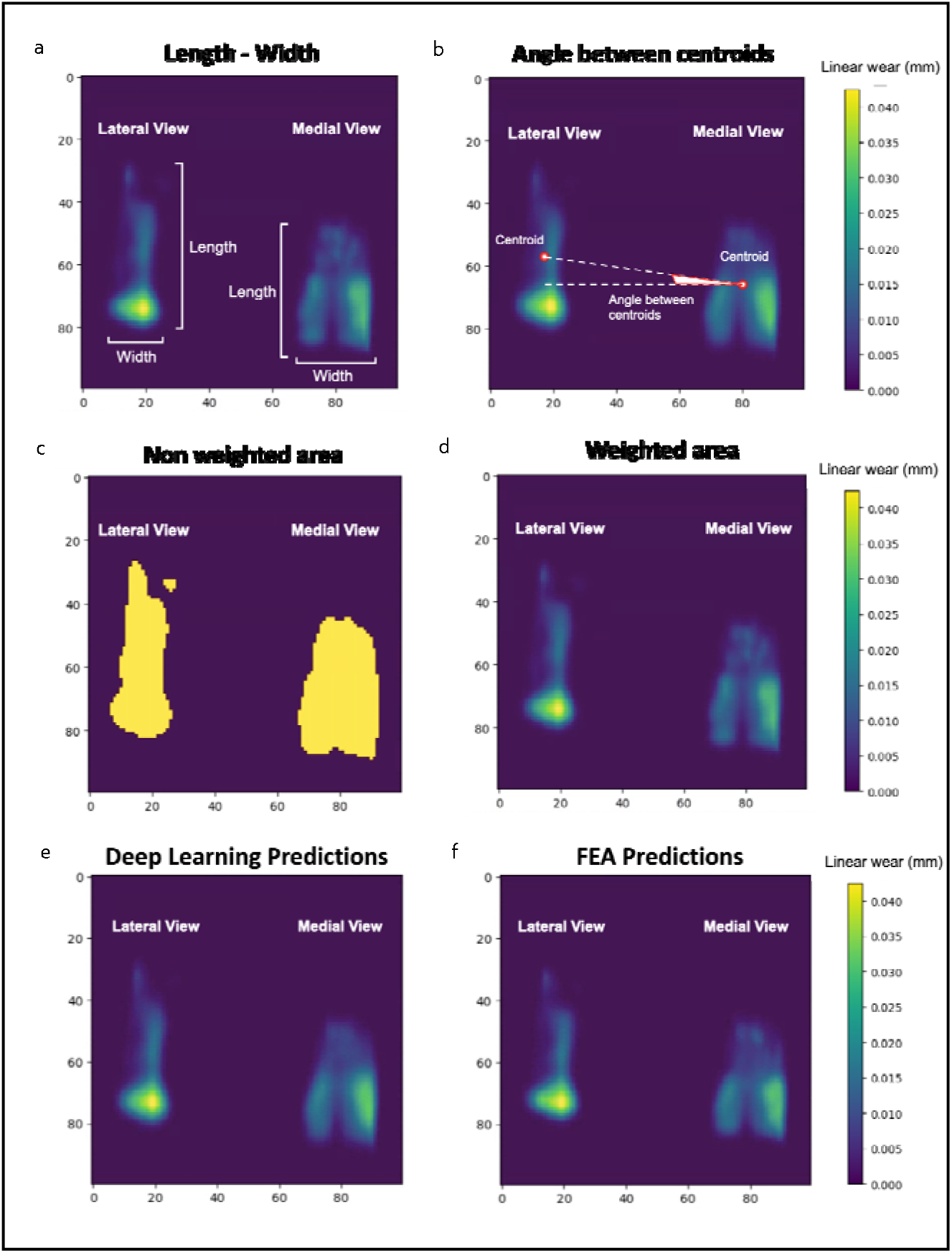
Figures 2.a through 2.d display the metrics utilized in this study, including the length and width of the wear maps (2.a), the angle between centroids (2.b), the non-weighted area (2.c), and the weighted area (2.d). Figures 2.e and 2.f illustrate the predictions from the deep learning and FEA models for a test set sample, respectively. Results for both the lateral and medial compartments are reported in these figures

## Results

Across these configurations, MAPE for the non-weighted wear scar area remained approximately 4% for lateral (sd: 0.28-0.43%) and medial compartment (sd: 0.38-0.49%), while for the weighted wear scar areas, MAPE values were approximately 12% for both compartments (sd lateral: 1.34-1.43%; sd medial: 1.05-1.13%). The MAPE for the angle between centroids was around 13% (sd: 0.89-1.42%) and 15% (sd: 1.15-1.47%), respectively considering centroids of the weighted and non-weighted area. Width and length measurements of the wear scar showed errors consistently below 6% (sd: 0.29-0.47%) for both compartments, except for the width of the lateral compartment at a 100×100 grid size, which had an error of 7.69% (sd: 0.77%) (**Table 2**). Additionally, Structural Similarity Index Measure (SSIM) values remained above 0.88 and Normalized Mutual Information (NMI) values above 0.89 for both compartments (**Table 3**). The model training times for grid sizes of 100×100, 150×150, and 200×200 were 672 seconds, 716 seconds, and 946 seconds, respectively, while inference times were less than two seconds for all configurations.

**Table 2:** Mean and standard deviation of absolute percentage error (APE) across different interpolation grids (100×100, 150×150, and 200×200). The comparison includes metrics for wear scar width, length, area, and angle between centroids (both non-weighted and weighted) for both lateral and medial compartments.

| Parameter | Absolute Percentage Error (APE) – 100x100 |  | Absolute Percentage Error (APE) – 150x150 |  | Absolute Percentage Error (APE) – 200x200 |  |
| --- | --- | --- | --- | --- | --- | --- |
|  | Mean | Std dev | Mean | Std dev | Mean | Std dev |
| Width (Lateral) | 7.69% | 0.77% | 3.57% | 0.39% | 5.58% | 0.47% |
| Length (Lateral) | 5.28% | 0.46% | 5.06% | 0.46% | 4.69% | 0.41% |
| Area – non-weighted (Lateral) | 3.51% | 0.43% | 4.05% | 0.37% | 3.42% | 0.28% |
| Area – weighted (Lateral) | 12.33% | 1.34% | 12.46% | 1.40% | 13.29% | 1.43% |
| Width (Medial) | 1.93% | 0.29% | 2.52% | 0.29% | 1.43% | 0.16% |
| Length (Medial) | 4.37% | 0.41% | 4.31% | 0.40% | 3.90% | 0.34% |
| Area – non-weighted (Medial) | 4.75% | 0.49% | 4.61% | 0.38% | 4.80% | 0.42% |
| Area – weighted (Medial) | 12.54% | 1.07% | 11.66% | 1.05% | 13.46% | 1.13% |
| Angle between centroids – non-weighted | 15.12% | 1.47% | 13.14% | 1.15% | 13.78% | 1.28% |
| Angle between centroids – weighted | 11.58% | 1.42% | 13.65% | 1.41% | 9.41% | 0.89% |

**Table 3:** Structural similarity Index measure (SSIM) and Normalized Mutual Information (NMI) values for each grid size with confidence intervals.

| Parameter | Interpolation grid: 100x100 | Interpolation grid: 150x150 | Interpolation grid: 200x200 |
| --- | --- | --- | --- |
| SSIM (lateral) | 0.89 (0.88, 0.90) | 0.90 (0.89, 0.91) | 0.95 (0.94, 0.96) |
| SSIM (medial) | 0.88 (0.87, 0.89) | 0.94 (0.94, 0.95) | 0.96 (0.95, 0.96) |
| NMI (lateral) | 0.94 (0.94, 0.95) | 0.95 (0.94, 0.95) | 0.89 (0.88, 0.91) |
| NMI (medial) | 0.96 (0.96, 0.97) | 0.96 (0.95, 0.96) | 0.96 (0.95, 0.96) |

## Discussions

We developed a surrogate modeling framework to predict TKR component wear using a deep learning model trained on FEA simulations. The deep learning model requires only kinematic and kinetic variables as input data, reducing the resources and time to calculate TKR component wear. Specifically, all input variables can be acquired using existing motion analysis techniques via wearable sensors, cameras or force plates^34,35,36^, removing the dependence on invasive methods such as embedded sensors within the implant^37,38,39^ or from complex musculoskeletal models^40,41,42^. With the rise of markerless motion capture and portable force plates, all input data can be easily obtained in a non-invasive manner in a clinical setting^43,43,45^.

This novel approach may have exciting future clinical implications to help identify patients at higher risk of implant failure. Markerless gait analysis workflows could measure the necessary kinematic and kinetics in the clinic and then be used to generate a resulting wear pattern^46^. This enhanced patient monitoring approach could help extend the lifespan of implants and improve patient outcomes. In this context, the role of the deep learning model is to capture the impact of gait patterns on wear distributions within the knee implant, which is a relationship that has been underscored in several studies^11,12^, and may help promote timely intervention to address gait kinematics and kinetics, which are modifiable with rehabilitation interventions and training.

As this is the first study utilizing deep learning to predict wear distribution in total joint replacement settings, comparing our outcomes with existing studies is challenging. Nonetheless, analyzing the differences in metrics between the lateral and medial compartments still remains informative. **Table 2** shows that for all grid setups, MAPE for the lateral compartment’s width consistently exceeds that of the medial compartment. This is likely due to lower values of wear scar width for the lateral compartment, where even minor errors result in a significant relative increase in MAPE. MAPE for the length of the wear scar is comparable between the two compartments, while the angle between centroids is slightly greater for the non-weighted compared to the weighted wear scar. It is worth noting that such angles are quite small, with the average angle between centroids being respectively around 5° and 10° for non-weighted and weighted wear scar. Therefore, errors around 15% in terms of MAPE correspond to slightly less than 1° for non-weighted and slightly more than 1° for weighted wear scar, which may not be clinically relevant. MAPE values for both non-weighted and weighted area of the wear scar are consistent across the compartments. However, the error for the weighted area is approximately three times higher than that for area. These disparities must be linked to small differences in predicted linear wear between deep learning and FEA predictions rather than in the gross wear scar geometry, and may not represent meaningful differences. Therefore, we also considered metrics^32,33^ that specifically focus on heatmap patterns to directly assess the spatial fidelity between FEA and deep learning heatmaps (SSIM) and the shared information between the heatmaps predicted by these two methodologies (NMI). When evaluating both metrics, images were cropped to isolate either the lateral or medial compartment. This approach ensures that the metrics are not affected by regions of the heatmaps with zero values. Given that the values for both metrics ranged between 0.88 and 0.96, we can consider the predictive performance of the models to be satisfactory^32,33^.

The primary advantage of using deep learning over FEA is the reduction in computational time required to determine the output linear wear. Training of deep learning models required approximately 7 minutes for the 100×100 model and 11 minutes for the 200×200 model, and inference took just a few seconds on an A5000 GPU with 24 GB of VRAM. In contrast, FEA required significantly more time, taking approximately 8 hours to process each batch of up to 4 samples. With 314 samples in the dataset distributed across 79 batches, FEA required roughly 26 days on a workstation with two Xeon E5-2680 CPUs running at 2.40 GHz and 256GB of RAM. Although our proposed methodology has not been applied to a clinical population, based on the results of this study, the proposed surrogate modeling framework appears to be a viable approach for predicting wear patterns in clinical settings.

The current study is not without limitations. First, the proposed deep learning methodology has been applied to a synthetic dataset^10^ rather than on data from patients. Given the novelty of the current approach, we decided to test it on synthetic data generated through an extensively validated framework^10^ as a first attempt. Having achieved promising results, next steps will involve applying and evaluating this methodology on a clinical dataset^47^. Secondly, the absence of component alignment data represents another limitation of this study. This issue could be addressed in future research by generating our training dataset at different component alignments, and accounting for these additional inputs in our deep learning framework. Clinically, component alignment can be approximated from post-operative x-rays^48,49,50^. Thirdly, although we propose an alternative approach to FEA for predicting wear distribution in TKR settings, a well validated FEA model is still initially needed to generate a high-fidelity training set for the deep learning model. Finally, while our model currently only accounts for level walking, other motion tasks beyond gait should be investigated to ensure that our deep learning framework accurately predicts wear maps across different activities of daily living. Some studies have explored the effects of different motion tasks on wear in TKR^16,17^, and employing our deep learning framework to these various tasks will be important for a clinical application.

## Conclusions

The current study introduces a novel deep learning approach for predicting wear maps in TKR settings, utilizing kinetic and kinematic gait patterns. The dataset comprises synthetic data featuring variations of ISO14243-3(2014) parameters, such as anterior/posterior translation, internal/external rotation, flexion/extension, and axial loading time series. A transformer-CNN architecture was implemented to capture temporal dependencies within the input multivariate time series and to accurately reconstruct output image data. The deep learning model’s predictions of length, width, area, and angle between wear scar centroids were evaluated using MAPE, while SSIM and NMI were employed to assess the overall similarity of wear maps between the deep learning and FEA predictions. Future work will involve applying and evaluating this methodology on patient data.

## Data Availability

All data produced in the present study are available upon reasonable request to the authors

